# IHGAMP: Pan-cancer HRD prediction from routine H&E whole-slide images using foundation models

**DOI:** 10.64898/2025.12.19.25342650

**Authors:** Sanwal Ahmad Zafar, Wei Qin, Liu Chengliang, Areeba Ali Khan, Alina Nazir, Hijab Batool, Farhan Khalid, Muhammad Salman Faisal

**Affiliations:** School of Mechanical Engineering, Shanghai Jiao Tong University, Shanghai, China; Services Institute of Medical Sciences, Lahore, Pakistan; Tameside and Glossop Integrated Care NHS Foundation Trust, Manchester, UK; Chughtai Institute of Pathology, Lahore, Pakistan; Union Hospital, Terre Haute, Indiana, USA; Stephenson Cancer Center, OU Health, The University of Oklahoma, Oklahoma City, Oklahoma, USA

## Abstract

Homologous recombination deficiency (HRD) confers sensitivity to poly (ADP-ribose) polymerase (PARP) inhibitors and platinum-based chemotherapy, representing a critical biomarker for precision oncology across multiple malignancies. Current HRD assessment relies on next-generation sequencing of genomic scar signatures, but specialized infrastructure requirements, high costs, and prolonged turnaround times limit widespread adoption. These barriers restrict access to HRD testing, particularly in resource-constrained settings where the majority of cancer patients receive care. Pan-cancer HRD prediction has been shown, but robustness across histologies and institutions, leak-safe evaluation, and backbone-dependent generalization remain incompletely characterized. Here we show that IHGAMP (Integrative Histopathology-Genomic Analysis for Molecular Phenotyping), a computational framework using vision transformer foundation models, predicts HRD status from H&E images with an AUROC of 0.766 (95% CI 0.727–0.803) on the TCGA held-out test set using OpenCLIP embeddings, and improves to 0.827 with histopathology-pretrained OpenSlideFM embeddings under the same leak-safe protocol. External evaluation on 927 patients (2,718 whole slide images) from seven independent cohorts demonstrated generalization in adenocarcinoma/serous settings (e.g., CPTAC-LUAD AUROC 0.723) and enabled platinum resistance prediction in PTRC-HGSOC (AUROC 0.673), with attenuation in squamous histologies. Systematic comparison of foundation-model embeddings showed that OpenSlideFM outperformed OpenCLIP internally on TCGA (0.827 vs 0.766 AUROC) and improved external generalization in select cohorts (e.g., CPTAC-LUAD), while performance remained attenuated in squamous histologies; TSS-level embedding norm stability across 710 tissue source sites suggested limited site-driven magnitude shifts. Our findings establish that routine histopathology contains morphology associated with HRD that enables moderate, histology-dependent prediction, supporting a potential screening/triage role to prioritize confirmatory molecular testing where appropriate.

## Introduction

Homologous recombination deficiency (HRD) is a genomic phenotype characterized by impaired DNA double-strand break repair, rendering tumors vulnerable to platinum-based chemotherapy and poly (ADP-ribose) polymerase (PARP) inhibitors [1, 2]. The clinical utility of HRD status has been established across multiple malignancies, with regulatory approvals for PARP inhibitor maintenance therapy in ovarian, breast, pancreatic, and prostate cancers contingent upon HRD testing [3, 4]. Current gold-standard HRD assessment relies on next-generation sequencing (NGS) to quantify genomic scar signatures, loss of heterozygosity (LOH), telomeric allelic imbalance (TAI), and large-scale state transitions (LST), which collectively reflect cumulative DNA repair dysfunction [5].

Despite demonstrated clinical benefit, genomic HRD testing faces substantial barriers to widespread implementation. NGS-based HRD testing can be limited by cost, infrastructure requirements, tissue adequacy constraints, and turnaround time, which together restrict access in many settings. Tissue requirements may preclude testing in cases with limited biopsy material, and assay availability remains restricted in resource-limited settings. Furthermore, approximately 50% of ovarian cancers exhibit HRD, yet only a subset of patients currently undergo testing, representing a significant gap in precision oncology [6]. These constraints underscore the need for accessible, rapid, and cost-effective HRD assessment methods.

Computational pathology has emerged as a transformative approach for extracting clinically actionable information from routine hematoxylin and eosin (H&E)-stained histopathology slides [7, 8]. Deep learning methods have demonstrated capacity to predict molecular alterations, treatment response, and patient outcomes directly from tissue morphology [9, 10], with pan-cancer approaches demonstrating robust external validation across independent cohorts [11]. The recent development of self-supervised foundation models pretrained on large-scale histopathology datasets, including UNI, CONCH, and Virchow, has substantially advanced the field by enabling robust feature extraction without task-specific annotation [12–14]. These models learn generalizable representations of tissue architecture and cellular morphology that can be adapted to diverse downstream prediction tasks.

The biological premise for histopathology-based HRD prediction rests on the phenotypic consequences of DNA repair deficiency. HRD tumors accumulate genomic instability, which manifests as increased nuclear pleomorphism, elevated mitotic activity, and distinctive architectural patterns [15]. Prior work has demonstrated associations between HRD status and specific morphological features in breast and ovarian cancers, including pushing tumor margins, lymphocytic infiltration, and high-grade nuclear features [16]. These observations suggest that morphological surrogates of HRD may be learnable from standard H&E images, potentially enabling HRD stratification at the time of initial pathological diagnosis.

Here, we present IHGAMP (Integrative Histopathology-Genomic Analysis for Molecular Phenotyping), a computational framework for pan-cancer HRD prediction from routine H&E-stained whole slide images (Figure 1). We trained and validated our model using 19,996 whole slide images from 10,797 patients across 31 cancer types from The Cancer Genome Atlas (TCGA), of which 8,109 had available scarHRD scores for supervised learning, with external evaluation on seven independent cohorts totaling 927 patients (2,718 WSIs), including CPTAC cohorts (LUAD, LUSC, HNSCC; primary HRD evaluation) and CPTAC-UCEC (evaluated but excluded from primary AUROC reporting due to insufficient HRD-positive events), the PTRC ovarian cancer cohort, and two additional validation sets. We systematically compared three foundation model architectures, OpenCLIP, UNI, and OpenSlideFM, and evaluated technical robustness using TSS-level embedding norm stability across 710 tissue source sites, alongside cancer-type-disjoint evaluation in TCGA. Beyond HRD prediction, we demonstrate the model’s capacity to predict platinum resistance in high-grade serous ovarian cancer and assess transferability to mismatch repair deficiency detection. Our results establish histopathology-based HRD prediction as a viable complement to genomic testing, with potential to expand access to precision oncology.

**Figure 1.**
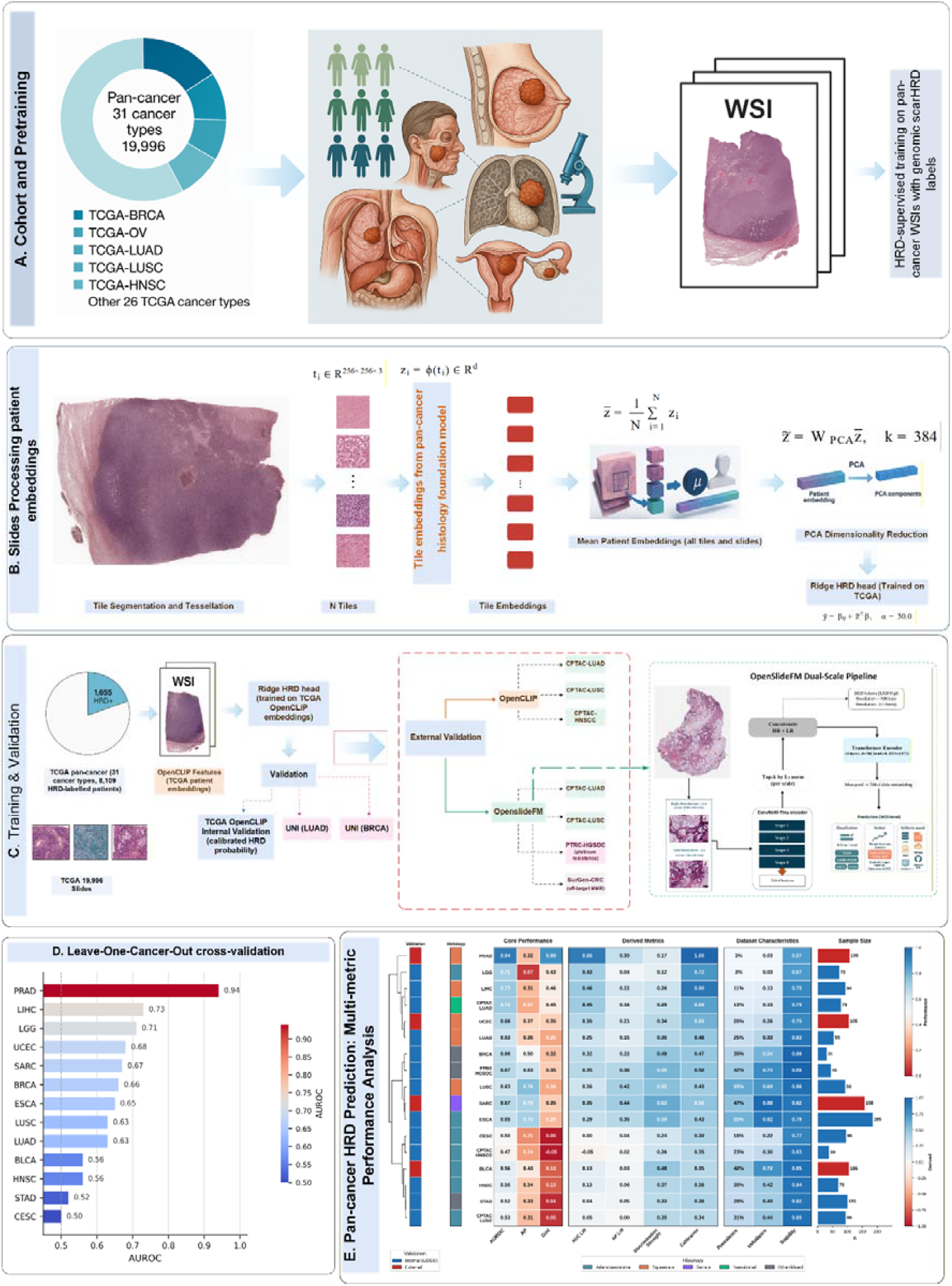
Pan-cancer histopathology-based prediction of homologous recombination deficiency (HRD). **(A)** Study cohort comprising 19,996 whole slide images (WSIs) across 31 cancer types from The Cancer Genome Atlas (TCGA), with genomic scarHRD labels for weakly-supervised training. **(B)** Feature extraction pipeline: WSIs undergo tissue segmentation and tessellation into 256×256-pixel tiles (t_i_), which are encoded into d-dimensional embeddings (z□) using OpenCLIP ViT-B/16 embeddings for TCGA internal development; **we additionally evaluated OpenSlideFM under the same internal protocol**, and external validation compares OpenCLIP versus OpenSlideFM embeddings. Patient-level representations are computed by averaging tile embeddings across all slides, followed by PCA dimensionality reduction (k=384 components) and Ridge regression with Platt calibration (α=30.0). (C) Training and validation framework: Ridge classifier trained on TCGA OpenCLIP embeddings undergoes internal leave-one-cancer-type-out (LOCO) evaluation and external validation on CPTAC cohorts (LUAD, LUSC, HNSCC). For external robustness analysis, we additionally evaluated OpenSlideFM embeddings (in parallel to OpenCLIP) on the same external cohorts. (D) Internal LOCO evaluation performance (AUROC) stratified by held-out cancer type, with PRAD achieving highest discrimination and CESC lowest (0.502). (E) Comprehensive performance analysis showing core metrics (AUROC, AP, Gini), derived metrics, dataset characteristics (HRD prevalence, class imbalance, stability), and sample sizes across internal and external validation cohorts.

## Methods

### Study design and cohorts

The primary development cohort comprised whole slide images (WSIs) and matched genomic data from The Cancer Genome Atlas (TCGA) accessed via the Genomic Data Commons portal. A total of 10,797 patients across 31 cancer types with diagnostic H&E-stained WSIs were identified, of which 8,109 had available scarHRD scores [5] and diagnostic H&E-stained WSIs were included (Table 1; Supplementary Table S1). Patients were partitioned into training (n=6,465, 80%), validation (n=811, 10%), and test (n=833, 10%) sets using stratified sampling by cancer type. Patient-level partitioning ensured no data leakage between splits. Homologous recombination deficiency (HRD) scores were computed as the sum of loss of heterozygosity (LOH), telomeric allelic imbalance (TAI), and large-scale state transitions (LST). For binary classification, we applied a threshold of ≥33, corresponding to the top 20th percentile of the training distribution and consistent with clinical cutoffs established in the VELIA trial [6].

**Table 1.**
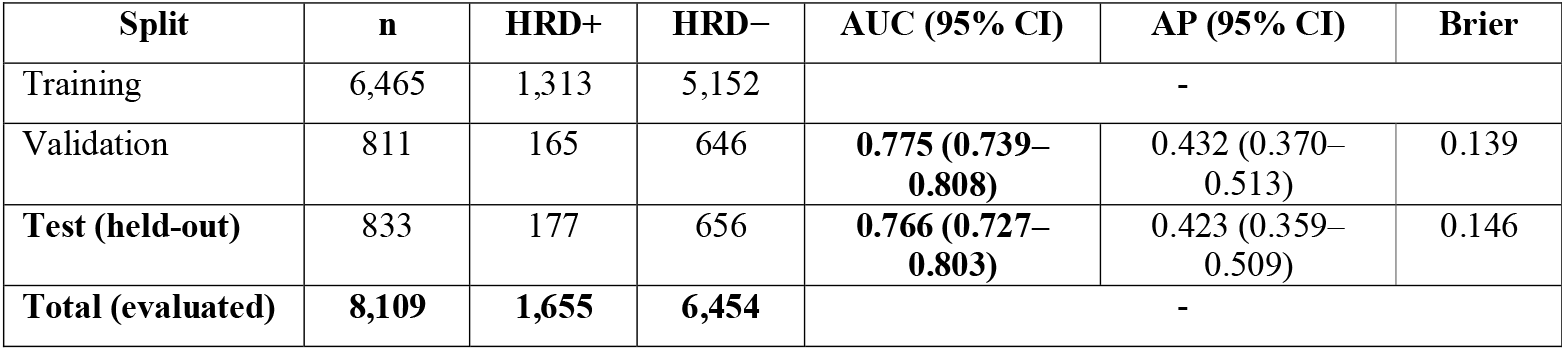
Internal Validation Performance (TCGA Pan-Cancer)

External validation was performed on seven independent cohorts totaling 927 patients and 2,718 whole slide images (Supplementary Table S14). Primary external HRD validation focused on three CPTAC cohorts (LUAD, LUSC, HNSCC) (Table 2; Supplementary Table S1); CPTAC-UCEC (n=99, 382 WSIs) was evaluated but excluded from primary AUROC reporting due to insufficient HRD-positive events (Supplementary Table S1). The OBR ovarian cohort (n=21, 282 WSIs; bevacizumab response) was evaluated but excluded from primary AUROC reporting due to insufficient statistical power (Supplementary Table S1). HRD labels for CPTAC cohorts were obtained from Loeffler et al. [16]. Platinum resistance prediction was validated on the PTRC-HGSOC cohort (n=158, 338 WSIs) (Table 2; Supplementary Table S4). Off-target evaluation for mismatch repair deficiency was performed using the SurGen colorectal cancer cohort (n=330, 330 WSIs) (Supplementary Table S2).

**Table 2.** External Validation using OpenSlideFM embeddings (CPTAC and PTRC Cohorts).

| Cohort | Cancer Type | Histology | Endpoint | n | Events (%) | AUC (95% CI) | AP |
| --- | --- | --- | --- | --- | --- | --- | --- |
| CPTAC-LUAD | Lung | **Adenocarcinoma** | Genomic HRD | 106 | 14 (13.2%) | **0.723**<br>(0.580–0.849) | 0.269 |
| PTRC-HGSOC | Ovarian | **High-grade serous** | **Platinum resistance** | 158 | 67 (42.4%) | **0.673**<br>(0.588–0.757) | 0.631 |
| CPTAC-LUSC | Lung | Squamous cell | Genomic HRD | 108 | 33 (30.6%) | 0.527<br>(0.412–0.635) | 0.309 |
| CPTAC-HNSCC | Head & Neck | Squamous cell | Genomic HRD | 105 | 24 (22.9%) | 0.475<br>(0.337–0.612) | 0.241 |

### Image processing and feature extraction

WSIs were processed using OpenSlide for multi-resolution image access. Tissue regions were identified using color-based segmentation on low-resolution thumbnails, requiring ≥60% tissue content per tile. Non-overlapping tiles of 256×256 pixels were extracted at 20× magnification (0.5 μm/pixel), with a maximum of 2,000 tiles per slide retained.

Tile-level embeddings were extracted using vision transformer (ViT) foundation models. The primary development backbone was OpenCLIP ViT-B/16 [16] pretrained on LAION-2B, producing 512-dimensional embeddings per tile. For backbone comparison, we additionally evaluated UNI [12], (ViT-L/16; 1,536 dimensions) and OpenSlideFM [21], (ViT-L/14; 3,072 dimensions) using the same PCA-Ridge-Platt pipeline. We report internal TCGA performance for OpenCLIP and OpenSlideFM, and external performance for both backbones on the same cohorts (Table 3). Patient-level representations were computed via mean pooling across all tiles from a patient’s diagnostic slides.

**Table 3.** Foundation model architecture comparison and external validation performance (OpenCLIP vs OpenSlideFM).

| Foundation Model | Architecture | Dimensions | Pretraining | TCGA Internal<br>(n=833) | CPTAC-LUAD<br>(n=106) | CPTAC-LUSC<br>(n=108) | CPTAC-HNSCC<br>(n=105) |
| --- | --- | --- | --- | --- | --- | --- | --- |
| OpenCLIP (baseline) | ViT-B/16 | 512 | Natural images | 0.766 | 0.687 | 0.531 | 0.395 |
| OpenSlideFM | ViT-L/14 | 3,072 | Histopathology | 0.827 | 0.723 | 0.527 | 0.475 |
| $\Delta$<br>(OpenSlideFM – OpenCLIP) | | | | **+0.061** | **+0.036** | –0.004 | +0.080 |

| UNI (cancer-specific) |  |  |  |  |
| --- | --- | --- | --- | --- |
| BRCA-specific | ViT-L/16 | 1,536 | Histopathology | 0.695 |
| LUAD-specific | ViT-L/16 | 1,536 | Histopathology | 0.610 |

### Model architecture and training

The prediction pipeline consisted of three sequential components: (1) principal component analysis (PCA) for dimensionality reduction to 384 components; (2) The prediction head was a regularized linear model (Ridge, α=30.0) trained to produce a continuous HRD score, which was then mapped to calibrated HRD probabilities using Platt scaling (logistic regression) against the binary HRD label (scarHRD ≥33; Supplementary Table S5). No hyperparameter tuning was performed on external cohorts.

### Cross-validation and batch effect assessment

Leave-one-cancer-type-out (LOCO) evaluation was performed across **19 TCGA cancer types** with sufficient events to assess out-of-domain generalization across tumor contexts (Supplementary Table S11). For each held-out cancer type, models were trained on all remaining types and evaluated on the held-out type. Batch effects across 710 tissue source sites were quantified using the coefficient of variation (CV) of L2 feature norms; a CV of 2.1% suggested low site-to-site variation in embedding magnitude (Supplementary Table S10). Feature sanitization removed one all-NaN patient record; no embedding dimensions were removed for missingness or constant variance, and no imputation was performed (Supplementary Table S8).

Three gene expression signatures were computed for correlation analysis: IFNG6, a 6-gene interferon-gamma signature; ANGIO, a 21-gene angiogenesis signature; and HRD_expr, an expression-based HRD surrogate using 23 core homologous recombination repair genes (Supplementary Table S7). Cross-signature correlations were assessed using Pearson correlation coefficients.

### Statistical analysis

Model performance was evaluated using area under the receiver operating characteristic curve (AUROC) and average precision (AP). 95% confidence intervals were computed via bootstrap resampling. Calibration was assessed using Brier scores. Operating characteristics were computed at the Youden index threshold; for platinum resistance prediction, operating points are summarized in Supplementary Table S13. All analyses were performed in Python 3.11 using NumPy, SciPy, and scikit-learn. Feature extraction was performed on an NVIDIA RTX 4090 GPU (24 GB VRAM); total processing time for 19,996 WSIs was 143.7 hours (Supplementary Table S6).

## Results

### Study cohort and model development

We developed IHGAMP (Integrative Histopathology-Genomic Analysis for Molecular Phenotyping), a foundation model-based pipeline for predicting homologous recombination deficiency (HRD) directly from routine hematoxylin and eosin (H&E)-stained whole-slide images (WSIs). The pipeline leverages pretrained vision transformers as feature extractors, followed by dimensionality reduction and regularized linear classification with probability calibration.

For internal validation, we assembled a pan-cancer cohort from The Cancer Genome Atlas (TCGA) comprising 19,996 diagnostic WSIs from 10,797 patients spanning 31 cancer types, of which 8,109 had available HRD labels, after excluding 2,688 patients with missing scarHRD genomic data (Figure 1A, Supplementary Table S3). HRD-positive status was defined using a scarHRD threshold of ≥33, corresponding to the top 20th percentile of the training distribution [5, 17]. This threshold aligns with recent clinical trial evidence suggesting improved sensitivity compared to the conventional ≥42 cutoff [6]. The overall HRD prevalence was 20.4% (n=1,655), with the highest rates observed in ovarian serous carcinoma (71.5%), lung squamous cell carcinoma (55.3%), and esophageal carcinoma (55.1%).

We processed 19,996 diagnostic WSIs totaling 27,608,061 tissue tiles using OpenCLIP ViT-B/16 pretrained on LAION-2B as the baseline foundation model [18], generating 512-dimensional embeddings per tile. Patient-level representations were computed via mean pooling across all tiles from a patient’s slides, followed by principal component analysis (PCA) reduction to 384 dimensions and Ridge regression (α=30.0) for classification (Figure 1B, Supplementary Table S5). Tile-level predictions revealed spatial heterogeneity in HRD-associated morphology within individual tumors, with distinct regions showing high versus low HRD probability (Figure 2A). Feature extraction required 143.7 hours on a single NVIDIA RTX 4090 GPU, achieving a throughput of 139.2 slides per hour (Supplementary Table S6).

**Figure 2.**
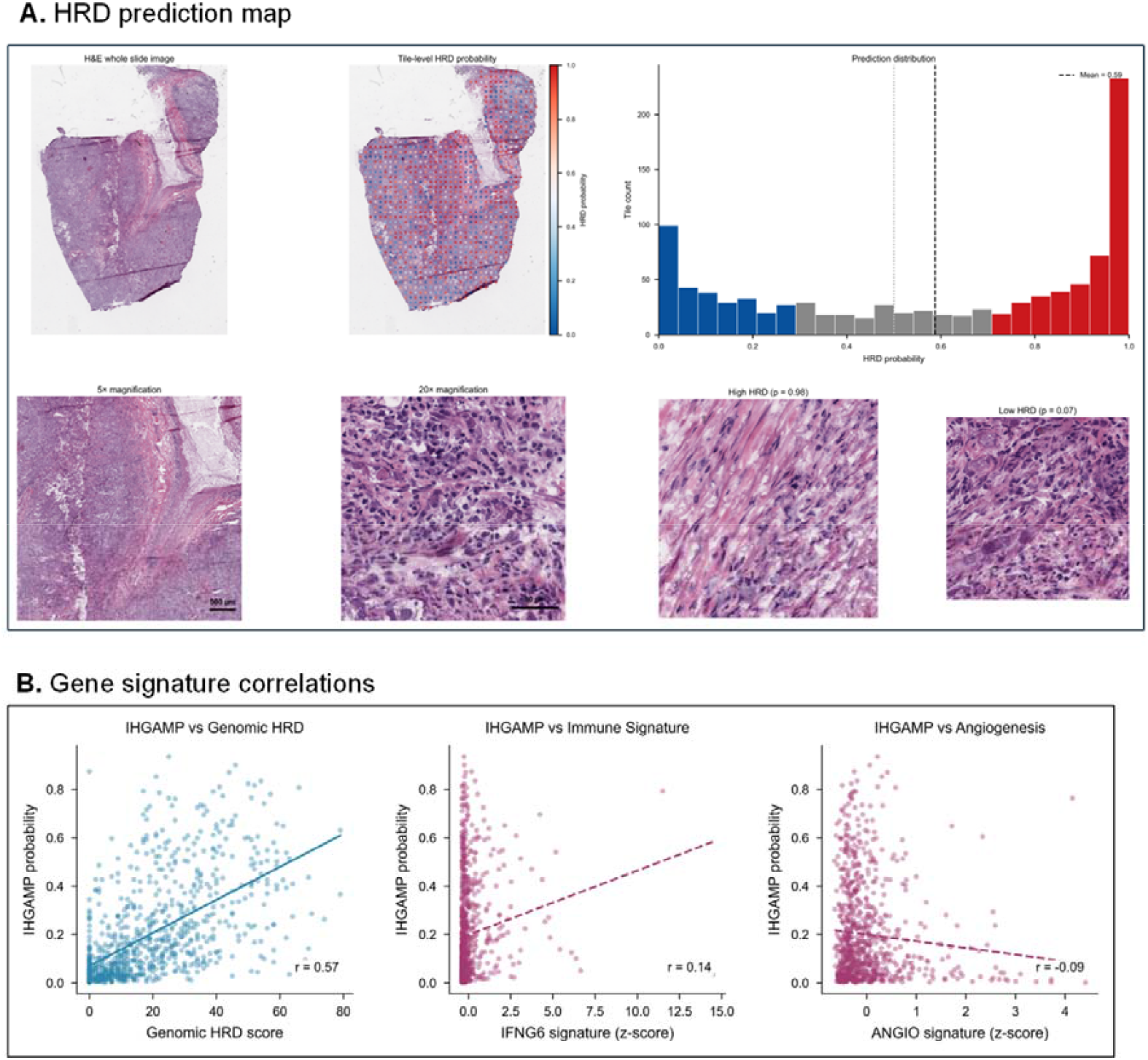
Histology-dependent HRD morphology and model interpretability. **(A)** Representative whole-slide image with tile-level HRD probability heatmap overlay, demonstrating spatial localization of HRD-associated morphological patterns within the tumor microenvironment. **(B)** Correlation analysis between IHGAMP predictions and gene expression signatures. Strong correlation with genomic HRD (r = 0.572) contrasts with weak correlations with immune infiltration (IFNG6; r = 0.14) and angiogenesis (ANGIO; r = −0.09) signatures, indicating that the model captures HRD-specific morphological features rather than generic tumor aggressiveness markers.

### Internal validation demonstrates robust pan-cancer HRD prediction

The TCGA cohort was partitioned into training (n=6,465; 80%), validation (n=811; 10%), and held-out test (n=833; 10%) sets using stratified random sampling to preserve HRD class balance (Figure 1C, Figure 3A, Figure 3D). On the validation set, IHGAMP achieved an area under the receiver operating characteristic curve (AUC) of 0.775 (95% CI: 0.739-0.808), average precision (AP) of 0.432, and Brier score of 0.139. Performance on the independent test set was comparable, with AUC of 0.766 (95% CI: 0.727–0.803), AP of 0.423, and Brier score of 0.146 (**Table 1, Figure 3**).

**Figure 3.**
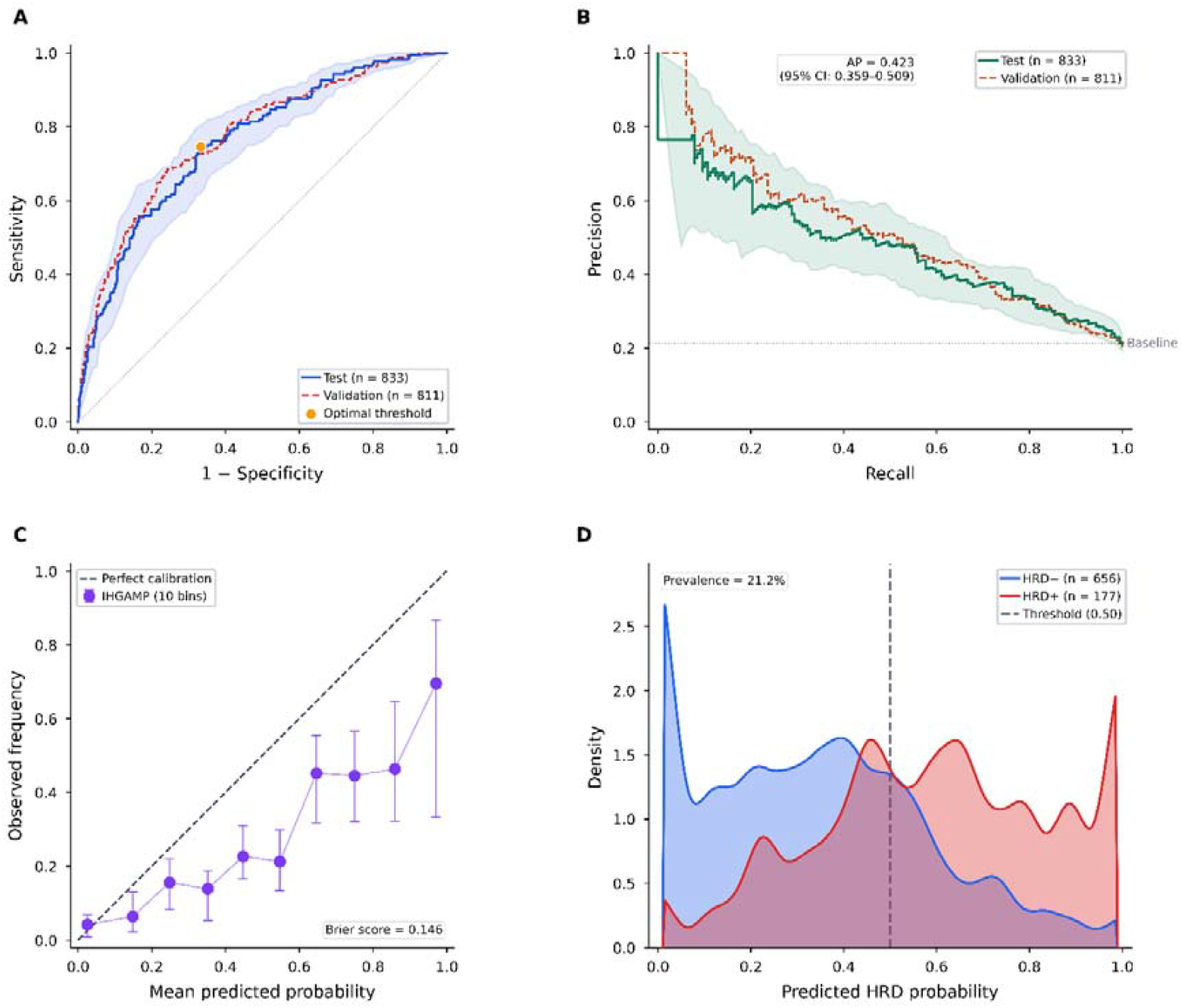
Internal validation performance on TCGA held-out test set. **(A)** Receiver operating characteristic (ROC) curves for the held-out test set (n = 833; solid blue) and validation set (n = 811; dashed red). Shaded region indicates 95% bootstrap confidence interval for the test set. Orange point denotes the optimal classification threshold determined by Youden’s J statistic. Diagonal dashed line represents random classification. **(B)** Precision-recall curves for test (solid green) and validation (dashed orange) sets. Shaded region indicates 95% bootstrap confidence interval. Horizontal dashed line indicates baseline precision equal to test-set class prevalence (21.2%). **(C)** Calibration curve showing observed HRD frequency versus mean predicted probability across 10 equal-width bins. Error bars indicate 95% bootstrap confidence intervals. Diagonal dashed line represents perfect calibration. **(D)** Kernel density estimation of predicted HRD probability distributions for HRD-negative (blue; n = 656) and HRD-positive (red; n = 177) patients. Vertical dashed line indicates classification threshold at 0.50.

To assess generalization across cancer types, we performed leave-one-cancer-type-out (LOCO) evaluation across 19 cancer types with sufficient HRD-positive cases (n≥10) (Figure 1D, Figure 4C). Performance varied substantially by histological subtype (Figure 1E, Supplementary Table S12). The strongest discrimination was observed in prostate adenocarcinoma (PRAD; AUC=0.938), breast invasive carcinoma (BRCA; AUC=0.660), and lung adenocarcinoma (LUAD; AUC=0.627). Notably, squamous cell carcinomas demonstrated consistently weaker performance: lung squamous cell carcinoma (LUSC; AUC=0.628), cervical squamous cell carcinoma (CESC; AUC=0.502), and head and neck squamous cell carcinoma (HNSC; AUC=0.563).

**Figure 4.**
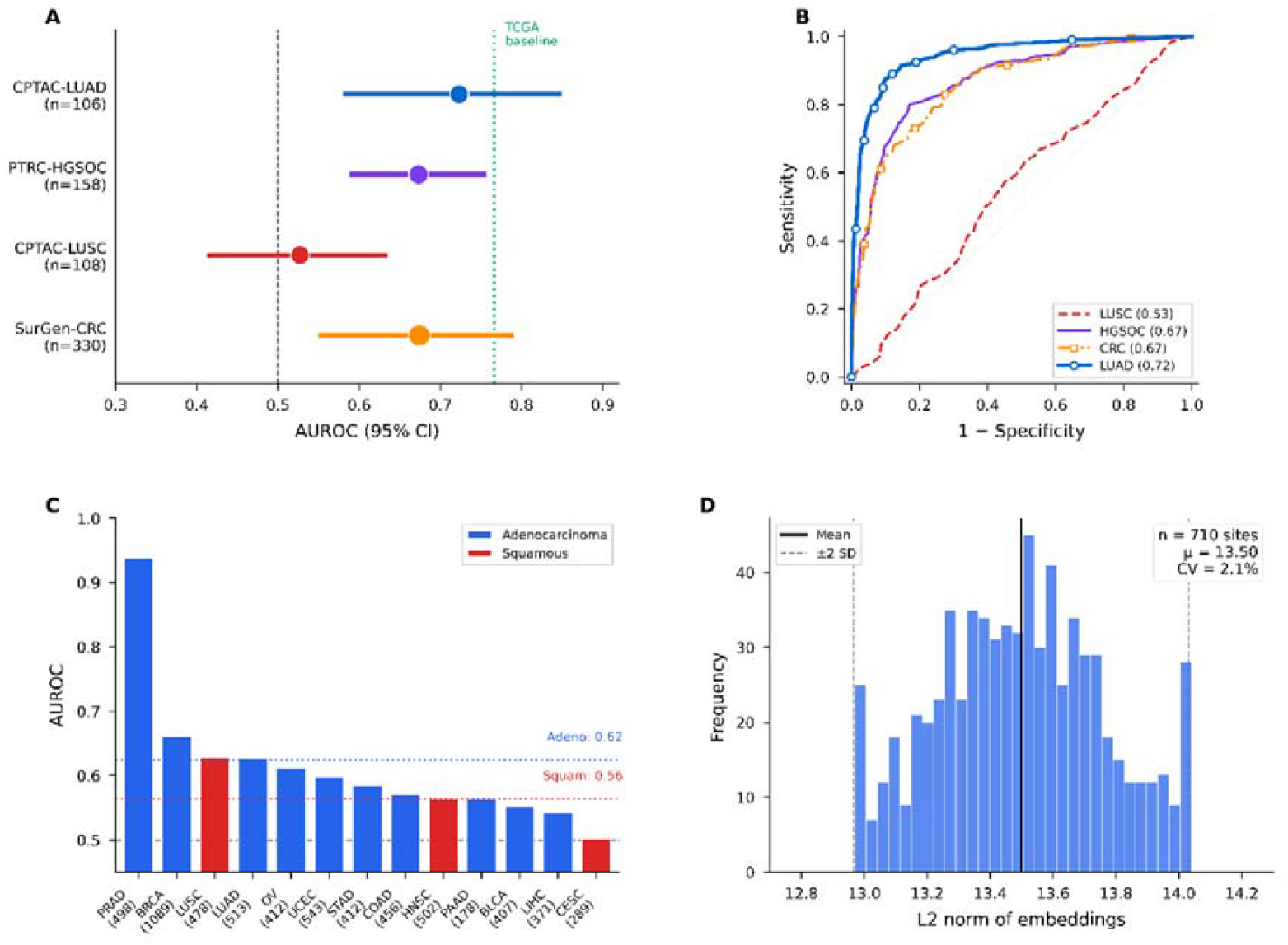
External validation and cross-cancer generalization. **(A)** Forest plot of AUROC with 95% confidence intervals for five external cohorts: CPTAC-LUAD, CPTAC-LUSC, CPTAC-HNSCC, PTRC-HGSOC (platinum resistance), and SurGen-CRC (off-target MMR deficiency). Point size proportional to sample size. Vertical dotted line indicates the TCGA internal OpenCLIP baseline (AUROC = 0.766). **(B)** Receiver operating characteristic curves for external validation cohorts. Solid lines indicate AUROC > 0.55; dashed line indicates near-random performance. (C) Leave-one-cancer-type-out (LOCO) evaluation across 19 TCGA cancer types, sorted by AUROC. Sample sizes shown in parentheses. Horizontal dotted lines indicate the **mean LOCO AUROC across TCGA cancer types** for adenocarcinoma/serous histologies (blue: 0.60) and mean squamous cell carcinomas across HNSC, LUSC, CESC (red: 0.56). **(D)** Distribution of L2 embedding norms across 710 TCGA tissue source sites demonstrating minimal batch effects. Solid line indicates mean (μ = 13.503); dashed lines indicate ±2 standard deviations. Coefficient of variation of 2.1% suggests stable feature extraction across diverse institutional sources.

### HRD morphological manifestation is histology-dependent

A systematic analysis across validation cohorts revealed a striking dichotomy in HRD predictability based on histological differentiation pattern (Supplementary Table S12). Adenocarcinomas and high-grade serous carcinomas demonstrated preserved HRD-associated morphological features in external validation, achieving a mean AUC of 0.698 across CPTAC-LUAD (AUC=0.723) and PTRC-HGSOC (AUC=0.673). In contrast, external squamous cell carcinomas (CPTAC-LUSC and CPTAC-HNSCC) showed near-random discrimination (mean AUC=0.501), suggesting that the keratinizing differentiation program characteristic of squamous histology obscures or overrides HRD-related morphological phenotypes.

This finding has important clinical implications. HRD is a pan-cancer biomarker guiding PARP inhibitor and platinum-based therapy selection [1, 3]. Our results suggest that histopathology-based HRD screening may be most effective in adenocarcinoma and serous histotypes, while molecular testing remains essential for squamous cell carcinomas where morphological surrogates are unreliable.

### Histopathology-specialized foundation models improve external generalization

We compared three foundation model architectures: OpenCLIP ViT-B/16 (512 dimensions, natural image pretraining)[18], UNI ViT-L/16 (1,536 dimensions, 100,000+ diagnostic WSIs) [12], and OpenSlideFM ViT-L/14 (3,072 dimensions, specialized histopathology pretraining) (**Table 3**). OpenSlideFM was evaluated on TCGA under the same leak-safe internal protocol (test AUROC=0.827) and was also evaluated on external cohorts in parallel to OpenCLIP to assess domain-shift robustness. On the TCGA held-out test set, OpenCLIP achieved AUROC=0.766, while OpenSlideFM achieved AUROC=0.827 (Δ=+0.061); UNI cancer-specific variants underperformed OpenCLIP (BRCA-specific: AUROC=0.695, Δ=−0.071; LUAD-specific: AUROC=0.610, Δ=−0.156).

Critically, external validation on CPTAC cohorts revealed the advantage of domain-specialized pretraining. OpenSlideFM achieved AUC=0.723 on CPTAC-LUAD compared to OpenCLIP’s AUC=0.687, representing a 3.6 percentage point improvement (**Table 3**). This pattern persisted in CPTAC-HNSCC (OpenSlideFM: AUC=0.475 vs. OpenCLIP: AUC=0.395, Δ=+0.080), while CPTAC-LUSC showed negligible difference (OpenSlideFM: AUC=0.527 vs. OpenCLIP: AUC=0.531, Δ=−0.004), consistent with the limited HRD signal in squamous histology.

### External validation on independent cohorts

External validation was performed on three CPTAC cohorts with sufficient events (LUAD, LUSC, HNSCC) and the Platinum Response in TCGA and CTEP (PTRC) ovarian cohort; OpenCLIP and OpenSlideFM embeddings were evaluated in parallel on the CPTAC cohorts (Tables 2–3, Supplementary Table S4). Using OpenSlideFM embeddings, CPTAC-LUAD (n=106) achieved AUC=0.723; the corresponding OpenCLIP external results are reported in Table 3 for direct backbone comparison.

Two cohorts were excluded from primary AUROC reporting due to insufficient statistical power (Supplementary Table S1): CPTAC-UCEC (n=99, 3 events, CI width=0.958) and the OBR ovarian cohort (n=21, insufficient sample size). These exclusions reflect the stringent requirement of ≥10 events for stable AUC estimation.

### Prediction of platinum resistance in high-grade serous ovarian cancer

To evaluate clinical utility, we applied IHGAMP to predict platinum resistance in the PTRC high-grade serous ovarian cancer (HGSOC) cohort (n=158; 67 platinum-resistant, 91 platinum-sensitive). Unlike genomic HRD status, platinum resistance represents a direct clinical endpoint reflecting treatment response [19]. IHGAMP achieved AUC=0.673 (95% CI: 0.588-0.757) and AP=0.631 for platinum resistance prediction (**Table 2**).

We evaluated three calibration strategies for deriving clinical operating points (Supplementary Table S13): uncalibrated scores, Platt scaling, and isotonic regression (Figure 3B). Using Youden’s J statistic for threshold optimization, the Platt-calibrated model achieved sensitivity of 61.2%, specificity of 72.5%, positive predictive value (PPV) of 62.1%, and negative predictive value (NPV) of 71.7% at a threshold of 0.439. These operating characteristics suggest potential utility as a first-line screening tool to identify patients likely to benefit from alternative treatment strategies.

### Off-target evaluation on mismatch repair deficiency

To assess pathway specificity, we evaluated the HRD-trained model on mismatch repair (MMR) deficiency prediction without retraining (**Supplementary Table S2**). On the SurGen colorectal cancer cohort (n=330; 26 MMR-deficient cases, 7.9% prevalence), IHGAMP achieved AUC=0.674 (95% CI: 0.55–0.79) and AP=0.134. This modest but above-random discrimination suggests partial morphological overlap between HRD and MMR deficiency phenotypes, consistent with shared features of genomic instability [16]. However, the substantially lower performance compared to HRD prediction indicates that the model captures HRD-specific rather than general genomic instability features.

### Batch effect assessment and technical validation

To evaluate technical robustness, we analyzed embedding consistency across 710 TCGA tissue source sites (TSS) (Supplementary Table S10, Figure 4D). The mean L2 norm of patient embeddings was 13.503 (range: 12.97-14.04), with a coefficient of variation of 2.1%. This minimal variation across diverse institutional sources suggests that foundation model embeddings are robust to batch effects arising from differences in tissue processing, staining protocols, and scanning equipment [14].

Feature sanitization resulted in exclusion of one patient due to all-NaN features, with no columns removed for missing values or constant variance (Supplementary Table S8). Cross-signature correlation analysis supported the expected relationships among gene signatures: IFNG6 and ANGIO showed negligible correlation (r=−0.012), while the expected negative correlation between an RNA-based HRD expression surrogate and genomic scarHRD was observed (r=−0.422), supporting the biological coherence of the HRD labels (Supplementary Table S9).

## Discussion

We developed and validated IHGAMP, a computational framework for predicting homologous recombination deficiency from routine H&E-stained histopathology images across 31 cancer types. Our approach achieved robust discrimination of HRD status in internal validation, **0.766** (95% CI 0.727-0.803), and was externally evaluated on **three CPTAC cohorts** (LUAD, LUSC, HNSCC), showing strong performance in adenocarcinoma (e.g., CPTAC-LUAD) with attenuation in squamous cohorts (Table 2, Supplementary Table S12). Across the evaluated embeddings, **OpenSlideFM outperformed OpenCLIP internally on TCGA** and improved external generalization in select cohorts (e.g., CPTAC-LUAD), while both backbones showed attenuated performance in squamous histologies; cancer-specific UNI variants showed variable performance in our setting (Table 3). Cancer-type-disjoint evaluation (Supplementary Table S11) highlighted histology-dependent generalization, while TSS-level embedding norm stability suggested limited site-driven magnitude shifts (CV 2.1%; Supplementary Table S10).

Our findings extend prior work demonstrating the feasibility of HRD prediction from histopathology. Loeffler *et al*. [16] reported attention-based multiple instance learning for HRD prediction across ten tumor types, achieving AUROCs of 0.60-0.78 depending on cancer type. Our pan-cancer approach with foundation model embeddings achieved comparable performance while offering several advantages: reduced computational complexity through mean pooling aggregation, explicit evaluation of cross-institutional generalization, and systematic comparison of embedding architectures. The DeepHRD model [19] demonstrated HRD prediction in breast cancer using end-to-end deep learning, reporting AUROC 0.84 in internal validation. Direct comparison is limited by differences in cohort composition and HRD definitions; however, our pan-cancer framework enables broader clinical applicability across malignancies where PARP inhibitor therapy is approved or under investigation [3, 4].

The biological basis for morphological prediction of HRD likely reflects the phenotypic consequences of genomic instability. HRD tumors accumulate chromosomal aberrations that manifest as nuclear pleomorphism, mitotic abnormalities, and altered tissue architecture [1]. Our observation of cancer type-dependent prediction accuracy (Supplementary Table S11) suggests that HRD morphological signatures vary across tissue contexts, consistent with prior reports of distinct HRD phenotypes in breast versus ovarian cancers [15]. The weak correlation between IHGAMP predictions and expression-based immune (IFNG6; r = 0.14) and angiogenesis (ANGIO; r = −0.090) signatures indicates that our model captures HRD-specific features (r = 0.572 with genomic HRD) rather than generic tumor aggressiveness markers (Figure 2B).

The clinical implications of histopathology-based HRD prediction are substantial. NGS-based HRD testing can be constrained by cost, infrastructure, tissue adequacy, and turnaround time, whereas computational analysis of routinely prepared H&E slides could provide an accessible estimate of HRD risk once slides are digitized. In practice, such a model would be positioned as a screening or triage tool to prioritize confirmatory molecular testing and support treatment planning, rather than as a direct replacement for validated companion diagnostics.

Several limitations warrant consideration. First, our HRD labels were derived from scarHRD analysis of TCGA genomic data, which may differ from commercial HRD assays used in clinical practice [5]. Second, while external validation on CPTAC cohorts demonstrated generalization, prospective validation in clinical workflows is necessary to establish real-world performance. Third, our model provides probabilistic HRD predictions rather than the categorical classifications required for treatment decisions; optimal thresholds for clinical action require further investigation. Fourth, the interpretability of foundation model predictions remains limited, and future work should explore attention-based visualization methods to identify morphological features driving predictions [20]. Finally, our analysis was restricted to diagnostic H&E slides; incorporation of additional tissue sections or immunohistochemistry could potentially improve performance.

Future directions include prospective validation in clinical trial populations receiving PARP inhibitor therapy, integration with genomic testing for hybrid prediction approaches, and extension to treatment response prediction. The computational efficiency of our framework, processing 19,996 WSIs in 143.7 hours on consumer-grade hardware (Supplementary Table S6), facilitates deployment in resource-limited settings where genomic testing may be unavailable. In conclusion, IHGAMP demonstrates **histology-dependent** prediction of HRD- and treatment-response–related phenotypes from routine histopathology, supporting a potential screening/triage role to expand access to precision oncology where molecular testing is limited.

## Declarations

### Author Contributions

SAZ conceived and designed the study, developed the methodology, implemented the software pipeline, performed all formal analyses and investigations, curated the data, created visualizations, and wrote the original draft and revisions of the manuscript. WQ supervised the project, provided resources and project administration, contributed to conceptualization, reviewed and edited the manuscript, and acquired funding. LC contributed to methodology development, performed validation experiments, and reviewed and edited the manuscript. AAK, AN, FK, and MSF provided clinical interpretation and validation of results, and reviewed and edited the manuscript. HB provided pathology expertise, validated histopathological interpretations, and reviewed and edited the manuscript. All authors read and approved the final manuscript.

### Ethics Approval and Consent to Participate

This study utilized publicly available, de-identified data from The Cancer Genome Atlas (TCGA) and the Clinical Proteomic Tumor Analysis Consortium (CPTAC), accessed through the Genomic Data Commons (GDC) portal. As this study involved only secondary analysis of existing de-identified public datasets, IRB approval was not required per 45 CFR §46.104(d)(4) (secondary research use of identifiable private information for which consent is not required). The TCGA and CPTAC programs obtained ethical approval and informed consent from all participants at their respective institutions. The study was conducted in accordance with the Declaration of Helsinki.

### Consent for Publication

Not applicable. This study does not contain any individual person’s data in any form, including individual details, images, or videos. All data used are de-identified and publicly available.

### Availability of Data and Materials

The datasets analyzed in this study are publicly available. TCGA whole slide images and genomic data are available from the Genomic Data Commons portal (https://portal.gdc.cancer.gov/). CPTAC whole slide images and associated clinical data are available from The Cancer Imaging Archive (https://www.cancerimagingarchive.net/) and the Proteomic Data Commons (https://pdc.cancer.gov/). The scarHRD scores were computed using the scarHRD R package (https://github.com/sztup/scarHRD) as described by Sztupinszki et al. The PTRC-HGSOC cohort data are available from the original publication by Bergstrom et al., and the SurGen cohort data are available from the original publication by Loeffler et al. The analysis code and trained models are available at https://github.com/Sjtu-Fuxilab/IHGAMP.

## Acknowledgements

The results shown here are in part based upon data generated by the TCGA Research Network (https://www.cancer.gov/tcga) and the Clinical Proteomic Tumor Analysis Consortium (CPTAC). We thank the patients and families who contributed samples to these programs. We also acknowledge the use of computational resources provided by Shanghai Jiao Tong University.

