## Supplementary Tables for "IHGAMP: Pan-cancer HRD prediction from routine H&E whole-slide images using foundation models"

### Supplementary Table S1. Excluded Cohorts Due to Insufficient Statistical Power

| **Cohort** | **Cancer Type** | **Endpoint** | **n** | **Events (%)** | **AUC** | **95% CI** | **CI Width** | **Exclusion Reason** |
| --- | --- | --- | --- | --- | --- | --- | --- | --- |
| CPTAC-UCEC | Endometrial | Genomic HRD | 99 | 3 (3.0%) | 0.594 | 0.021–0.979 | **0.958** | Insufficient events (n=3) |
| OBR | Ovarian | Bevacizumab | 21 | 12 (57.1%) | 0.852 | 0.546–1.000 | **0.454** | Insufficient sample size |

### Supplementary Table S2. Off-Target Evaluation on Mismatch Repair Deficiency

| **Cohort** | **Cancer Type** | **Target Pathway** | **Model Training** | **n** | **Events (%)** | **AUC (95% CI)** | **AP** | **Interpretation** |
| --- | --- | --- | --- | --- | --- | --- | --- | --- |
| SurGen | Colorectal | MMR deficiency | HRD (off-target) | 330 | 26 (7.9%) | 0.674 (0.55–0.79) | 0.134 | Partial overlap |

### Supplementary Table S3. Complete TCGA Cancer Type Distribution

| **Cancer** | **Full Name** | **n** | **HRD+** | **HRD−** | **% HRD+** |
| --- | --- | --- | --- | --- | --- |
| BRCA | Breast invasive carcinoma | 955 | 281 | 674 | 29.4% |
| LGG | Low-grade glioma | 498 | 8 | 490 | 1.6% |
| LUAD | Lung adenocarcinoma | 488 | 150 | 338 | 30.7% |
| HNSC | Head and neck squamous cell | 474 | 134 | 340 | 28.3% |
| LUSC | Lung squamous cell carcinoma | 459 | 254 | 205 | 55.3% |
| PRAD | Prostate adenocarcinoma | 458 | 18 | 440 | 3.9% |
| THCA | Thyroid carcinoma | 453 | 0 | 453 | 0.0% |
| BLCA | Bladder urothelial carcinoma | 391 | 150 | 241 | 38.4% |
| STAD | Stomach adenocarcinoma | 368 | 126 | 242 | 34.2% |
| SKCM | Skin cutaneous melanoma | 350 | 33 | 317 | 9.4% |
| LIHC | Liver hepatocellular carcinoma | 341 | 36 | 305 | 10.6% |
| KIRC | Kidney renal clear cell | 338 | 1 | 337 | 0.3% |
| CESC | Cervical squamous cell | 272 | 44 | 228 | 16.2% |
| KIRP | Kidney renal papillary cell | 265 | 1 | 264 | 0.4% |
| COAD | Colon adenocarcinoma | 257 | 11 | 246 | 4.3% |
| SARC | Sarcoma | 223 | 106 | 117 | 47.5% |
| UCEC | Uterine corpus endometrial | 174 | 42 | 132 | 24.1% |
| OV | Ovarian serous cystadenocarcinoma | 172 | 123 | 49 | 71.5% |
| PCPG | Pheochromocytoma/paraganglioma | 156 | 0 | 156 | 0.0% |
| ESCA | Esophageal carcinoma | 147 | 81 | 66 | 55.1% |
| PAAD | Pancreatic adenocarcinoma | 140 | 9 | 131 | 6.4% |
| GBM | Glioblastoma multiforme | 123 | 1 | 122 | 0.8% |
| THYM | Thymoma | 103 | 0 | 103 | 0.0% |
| READ | Rectum adenocarcinoma | 83 | 3 | 80 | 3.6% |
| MESO | Mesothelioma | 80 | 7 | 73 | 8.8% |
| UVM | Uveal melanoma | 78 | 0 | 78 | 0.0% |
| ACC | Adrenocortical carcinoma | 74 | 2 | 72 | 2.7% |
| KICH | Kidney chromophobe | 60 | 0 | 60 | 0.0% |
| UCS | Uterine carcinosarcoma | 56 | 29 | 27 | 51.8% |
| DLBC | Diffuse large B-cell lymphoma | 37 | 1 | 36 | 2.7% |
| CHOL | Cholangiocarcinoma | 36 | 4 | 32 | 11.1% |
| **Total** | 31 cancer types | **8,109** | **1,655** | **6,454** | **20.4%** |

### Supplementary Table S4. External Validation Overview

| **Cohort** | **Cancer Type** | **Histology** | **Endpoint** | **n** | **Events (%)** | **AUC (95% CI)** | **AP** | **Status / Reason** |
| --- | --- | --- | --- | --- | --- | --- | --- | --- |
| CPTAC-LUAD | Lung | Adenocarcinoma | Genomic HRD | 106 | 14 (13.2%) | **0.723 (0.580–0.849)** | 0.269 | **Included (main)** |
| PTRC-HGSOC | Ovarian | High-grade serous | Platinum resistance | 158 | 67 (42.4%) | **0.673 (0.588–0.757)** | 0.631 | **Included (main)** |
| CPTAC-LUSC | Lung | Squamous cell | Genomic HRD | 108 | 33 (30.6%) | 0.527 (0.412–0.635) | 0.309 | Included (main) |
| CPTAC-HNSCC | Head & Neck | Squamous cell | Genomic HRD | 105 | 24 (22.9%) | 0.475 (0.337–0.612) | 0.241 | Included (main) |
| CPTAC-UCEC | Endometrial | Mixed | Genomic HRD | 99 | 3 (3.0%) | 0.594 (0.021–0.979) | — | Excluded – Insufficient events (n=3) |
| OBR | Ovarian | Mixed | Bevacizumab response | 21 | 12 (57.1%) | 0.852 (0.546–1.000) | — | Excluded – Insufficient sample size |
| SurGen | Colorectal | Adenocarcinoma | MMR deficiency (off-target) | 330 | 26 (7.9%) | 0.674 (0.55–0.79) | 0.134 | Off-target evaluation |

### Supplementary Table S5. Model Hyperparameters

| **Parameter** | **Value** | **Justification** |
| --- | --- | --- |
| PCA components | 384 | Variance retention, grid search |
| Ridge α | 30.0 | RidgeCV optimization |
| HRD threshold (scarHRD) | ≥33.0 | Top-20% of training distribution |
| Bootstrap replicates (internal) | 200 | Computational efficiency |
| Bootstrap replicates (external) | 2,000 | Robust CI estimation |
| Random seed | 42 | Reproducibility |
| Train/Val/Test split | 80/10/10 | Stratified by cancer type |
| Platt calibration | Logistic regression | Probability calibration on validation set |
| K-fold CV (signature prediction) | 5 | Cross-validation for RNA surrogates |

### Supplementary Table S6. Computational Resources and Processing Statistics

| **Resource/Parameter** | **Value** |
| --- | --- |
| GPU | NVIDIA GeForce RTX 4090 (23.99 GB VRAM) |
| CUDA version | 12.1 |
| Mixed precision (AMP) | True |
| TF32 | Enabled |
| Total TCGA slides | 20,000 |
| Slides successfully embedded | 19,996 |
| Failed slides | 4 |
| Unique tissue source sites (TSS) | 710 |
| Patch size | 256 × 256 pixels |
| Patches per slide (cap) | 2,000 |
| Total tiles extracted | 27,608,061 |
| Stride | 384 pixels |
| Magnification level | Level 0 (highest resolution) |
| Preprocessing | GPU resize→224 + CLIP mean/std normalization |
| OpenCLIP backbone | ViT-B/16 (laion2b_s34b_b88k) |
| OpenCLIP embedding dim | **512** |
| OpenSlideFM embedding dim | **3,072 (ViT-L/14)** |
| UNI embedding dim | 1,536 (ViT-L/16) |
| Total processing time | 143.7 hours (517,320 seconds) |
| Throughput | 139.2 slides/hour, 53 tiles/second |
| Feature aggregation | Mean pooling (patient-level) |

### Supplementary Table S7. Gene Signature Panels

| **Signature** | **Genes** | **N** | **Source** |
| --- | --- | --- | --- |
| IFNG6 | IFNG, STAT1, IDO1, CXCL9, CXCL10, HLA-DRA | 6 | Ayers et al. |
| ANGIO | VEGFA, KDR, FLT1, ANGPT2, TEK, ENG, PECAM1, VWF, COL4A1, COL4A2, MMP2, MMP9, HIF1A, PDGFB, PDGFRB, ITGAV, ITGB3, ICAM1, SELE, PGF, ELN | 21 | Hallmark |
| HRR_CORE | BRCA1, BRCA2, PALB2, RAD51, RAD51C, RAD51D, BARD1, BRIP1, ATM, ATR, CHEK1, CHEK2, XRCC2, XRCC3, MRE11, RAD50, NBN, FANCA, FANCD2, FANCM, RPA1, RFC2, BLM | 23 | DDR pathway |

### Supplementary Table S8. Feature Sanitization Report

| **Sanitization Step** | **Result** |
| --- | --- |
| Input feature dimensions | 512 (OpenCLIP ViT-B/16) |
| Minimum non-NA column ratio | 0.98 |
| Minimum variance threshold | 1×10⁻¹² |
| Patients dropped (all-NaN) | **1** |
| Columns dropped (missing) | **0** |
| Columns dropped (constant) | **0** |
| Imputed cells | **0** |
| Final feature dimensions | **512 → 384 (after PCA)** |

### Supplementary Table S9. Cross-Signature Correlations

| **Signature Pair** | **Pearson r** | **Interpretation** |
| --- | --- | --- |
| IFNG6 ~ ANGIO | −0.012 | Negligible (independent axes) |
| IFNG6 ~ HRD_expr | −0.194 | Weak negative |
| ANGIO ~ HRD_expr | +0.160 | Weak positive |
| HRD_expr ~ HRD_genomic | **−0.422** | **Moderate negative (expected)** |

### Supplementary Table S10. Batch Effect Assessment (Tissue Source Sites)

| **Metric** | **Value** | **Range** | **Interpretation** |
| --- | --- | --- | --- |
| Total tissue source sites (TSS) | 710 | — | TCGA-wide |
| Total slides processed | 19,996 | — | — |
| Mean L2 norm (overall) | **13.503** | 12.97–14.04 | **Consistent** |
| L2 norm std dev | 0.287 | — | **Minimal** |
| L2 norm CV (%) | 2.1% | — | **Excellent** |
| Sites with >100 slides | 45 | — | — |
| Sites with 1 slide only | 267 | — | — |

### Supplementary Table S11. Leave-One-Site-Out Cross-Validation by Cancer Type

| **Site** | **Cancer Type** | **n** | **HRD+** | **HRD−** | **% HRD+** | **AUC** | **AP** | **Status** |
| --- | --- | --- | --- | --- | --- | --- | --- | --- |
| BRCA | Breast invasive carcinoma | 185 | 65 | 120 | 35.1% | 0.660 | 0.497 | Moderate |
| LGG | Low-grade glioma | 101 | 3 | 98 | 3.0% | **0.714** | 0.065 | Good* |
| LUAD | Lung adenocarcinoma | 96 | 24 | 72 | 25.0% | 0.627 | 0.359 | Moderate |
| PRAD | Prostate adenocarcinoma | 95 | 3 | 92 | 3.2% | **0.938** | 0.324 | Excellent* |
| HNSC | Head and neck squamous cell | 94 | 28 | 66 | 29.8% | 0.563 | 0.340 | Poor |
| LUSC | Lung squamous cell carcinoma | 93 | 55 | 38 | 59.1% | 0.628 | 0.762 | Moderate |
| BLCA | Bladder urothelial carcinoma | 79 | 33 | 46 | 41.8% | 0.564 | 0.435 | Poor |
| STAD | Stomach adenocarcinoma | 73 | 21 | 52 | 28.8% | 0.518 | 0.325 | Random |
| SKCM | Skin cutaneous melanoma | 72 | 7 | 65 | 9.7% | 0.490 | 0.108 | Random |
| LIHC | Liver hepatocellular carcinoma | 70 | 8 | 62 | 11.4% | **0.732** | 0.308 | Good |
| KIRC | Kidney renal clear cell | 69 | 0 | 69 | 0.0% | — | — | No events |
| CESC | Cervical squamous cell | 55 | 10 | 45 | 18.2% | 0.502 | 0.212 | Random |
| KIRP | Kidney renal papillary cell | 54 | 0 | 54 | 0.0% | — | — | No events |
| COAD | Colon adenocarcinoma | 53 | 3 | 50 | 5.7% | 0.473 | 0.207 | Below random* |
| SARC | Sarcoma | 45 | 21 | 24 | 46.7% | 0.673 | 0.702 | Moderate |
| UCEC | Uterine corpus endometrial | 39 | 8 | 31 | 20.5% | 0.677 | 0.368 | Moderate |
| PCPG | Pheochromocytoma/paraganglioma | 34 | 0 | 34 | 0.0% | — | — | No events |
| ESCA | Esophageal carcinoma | 31 | 17 | 14 | 54.8% | 0.647 | 0.704 | Moderate |
| PAAD | Pancreatic adenocarcinoma | 31 | 1 | 30 | 3.2% | **1.000** | 1.000 | Perfect* |

### Supplementary Table S12. Histology-Dependent HRD Morphological Manifestation

| **Histological Category** | **Validation Cohorts** | **Total n** | **Events** | **Mean AUC** | **TCGA LOCO** | **CPTAC/PTRC** |
| --- | --- | --- | --- | --- | --- | --- |
| **Adenocarcinoma / Serous** | LUAD, HGSOC | 264 | 81 | **0.674** | 0.627 (LUAD) | **0.673–0.723** |
| Squamous cell carcinoma | CPTAC-LUSC, CPTAC-HNSCC | 213 | 57 | 0.501 | 0.502–0.628 | 0.475–0.527 |

### Supplementary Table S13. Clinical Operating Points for Platinum Resistance Prediction (PTRC-HGSOC)

| **Calibration** | **Threshold** | **Sensitivity** | **Specificity** | **PPV** | **NPV** | **Balanced Acc** | **Pred Positive** |
| --- | --- | --- | --- | --- | --- | --- | --- |
| Uncalibrated | 0.505 | 0.642 | 0.681 | 0.597 | 0.721 | 0.662 | 72/158 |
| **Platt (recommended)** | 0.439 | **0.612** | **0.725** | 0.621 | 0.717 | **0.669** | 66/158 |
| Isotonic | 0.375 | 0.627 | 0.670 | 0.583 | 0.709 | 0.649 | 72/158 |

**Supplementary Table S14. External Validation Cohort Summary**

| Cohort | Patients | WSIs | Endpoint | Status |
| --- | --- | --- | --- | --- |
| CPTAC-LUAD | 106 | 492 | Genomic HRD | Main (Table 2) |
| CPTAC-LUSC | 108 | 473 | Genomic HRD | Main (Table 2) |
| CPTAC-HNSCC | 105 | 421 | Genomic HRD | Main (Table 2) |
| CPTAC-UCEC | 99 | 382 | Genomic HRD | Excluded (S1) |
| PTRC-HGSOC | 158 | 338 | Platinum resistance | Main (Table 2) |
| OBR | 21 | 282 | Bevacizumab response | Excluded (S1) |
| SurGen | 330 | 330 | MMR deficiency | Off-target (S2) |
| Total | **927** | **2,718** | — | — |
